# Bronchoalveolar Lavage Metagenomic Sequencing in the Early Post-Lung Transplantation Period: A Pilot Comparison with Microbiologic Culture

**DOI:** 10.64898/2026.09.17.26363318

**Authors:** Georgios D. Kitsios, Michael A. Sy, Xiaohong Wang, Andrew Craig, Alicia Rizzo, John R. Francisco, Shulin Qin, Matthew Hensley, Kailey Hughes Kramer, Kentaro Noda, Panayiotis Benos, Chadi Hage, Pablo Sanchez, Alison Morris, Ghady Haidar, Mark E. Snyder

## Abstract

Conventional bronchoalveolar lavage (BAL) culture has limited sensitivity in lung transplant recipients due to universal antimicrobial prophylaxis and inability to detect non-culturable organisms. We prospectively enrolled 19 adult lung transplant recipients undergoing serial bronchoscopies during the immediate post-transplant period. BAL samples (N=33) were tested in parallel with conventional culture, microbial cell-free DNA (mcfDNA) sequencing with quantitative analysis, and long-read sequencing, using a unified three-tier organism classification framework. BAL host-response biomarkers were profiled concurrently. mcfDNA sequencing detected pathogens in 19 of 33 episodes (58%) versus 8 of 33 by conventional culture (24%); organism-level concordance was low (Cohen’s kappa 0.18). In 16 episodes where mcfDNA detected a pathogen not recovered by culture, 8 across 3 participants represented predictive detections — the same organism subsequently confirmed as causing invasive infection 2–46 days later, including fatal extensively drug-resistant Pseudomonas pneumonia, vancomycin-resistant Enterococcal surgical site infection, and candidemia. Long-read sequencing had low yield due to contaminating human DNA. Quantitative mcfDNA burden correlated positively with multiple alveolar inflammatory biomarkers. BAL mcfDNA metagenomics identified clinically relevant pathogens not recovered by conventional culture, including predictive detections and non-culturable organisms, supporting a role for metagenomic surveillance in the early post-transplant period.

## INTRODUCTION

Lung transplant recipients face a disproportionately high burden of infectious complications compared to other solid organ transplant recipients, with lower respiratory tract infections (LRTIs) are a leading cause of morbidity and mortality in the first year after transplantation.^1–3^ Recipients are particularly vulnerable in the early post-transplant period: surgical disruption of the bronchial mucosa, loss of cough reflex and mucociliary clearance in the allograft, intense immunosuppression, and prolonged mechanical ventilation create a permissive environment for both donor-derived and nosocomial pathogens.^4^ Prompt and accurate microbiological diagnosis is central to post-transplant care, as delays in pathogen identification compromise the ability to de-escalate empiric broad-spectrum antimicrobials or target treatment to resistant organisms.^5^

Bronchoscopy with bronchoalveolar lavage (BAL) remains the cornerstone diagnostic procedure for suspected LRTI after lung transplantation.^6^ However, conventional culture-based BAL microbiology has well-recognized limitations in this population. Sensitivity is substantially reduced by prior or ongoing antimicrobial therapy, which is nearly universal given standard prophylaxis regimens. Fastidious and difficult-to-culture organisms, such as *Mycoplasma* or *Ureaplasma* species, and certain fungi, can be systematically missed by culture-based methods^1,6^. Turnaround times of 48–72 hours for bacterial cultures and up to two weeks for fungal and mycobacterial cultures delay definitive diagnosis and prolong empiric therapy.^1,6^ Furthermore, distinguishing true pathogens from colonizing or commensal organisms in the immunocompromised allograft remains a persistent interpretive challenge.^7^

Metagenomic sequencing offers a pathogen-agnostic alternative that bypasses many of these limitations by detecting nucleic acid from any organism present in a sample, regardless of viability, cultivability, or prior antibiotic exposure.^8–10^ Multiple metagenomic sequencing technologies (including short-read sequencing, long-read platforms, and microbial cell-free DNA (mcfDNA) approaches) have been applied to respiratory specimens with varying workflow requirements and turnaround times.^11,12^ Among these, two platforms have demonstrated particular potential for near-clinical deployment. The Karius Focus™ | BAL test is a commercially available mcfDNA sequencing assay that detects microbial DNA shed into airway lining fluid;^12,13^ by selectively processing the cell-free supernatant fraction, it mitigates the host-background signal that challenges whole cell metagenomic approaches and returns results within 24–48 hours.^14^ Oxford Nanopore Technology (ONT)-based sequencing offers an alternative approach, enabling real-time data generation at the point of sequencing and the potential for same-day pathogen identification without the infrastructure requirements of centralized laboratory platforms^15–17^. Both approaches have been evaluated in respiratory infection diagnostics, but comparisons of BAL metagenomics against conventional microbiology in the immediate post-lung transplant period are lacking.

The clinical relevance of metagenomic pathogen detection in the lung allograft remains to be established. To address this gap, we report a prospective cohort of lung transplant recipients undergoing serial bronchoscopies in the immediate post-transplant period to compare Karius BAL mcfDNA metagenomics and ONT long-read sequencing against conventional culture applied to the same BAL specimens. We concurrently profiled BAL biomarkers to characterize the alveolar host response in the context of metagenomic findings.

## METHODS

### Study design and participants

We conducted a prospective, single-center, observational study of adult lung transplant recipients at the University of Pittsburgh Medical Center (UPMC) as part of the KARL (Karius After Recent Lung transplant) cohort, as previously described.^18^ Thirty-one adult recipients were enrolled between April 2021 and December 2022 and followed through 6 months post-transplant. The present analysis focuses on 19 participants from whom excess BAL fluid aliquots were prospectively collected for research metagenomics testing. The study was approved by the University of Pittsburgh IRB (STUDY19080225) and all participants provided written informed consent.

### BAL sampling

Bronchoscopy was performed at the discretion of the clinical transplant team for standard indications, including scheduled surveillance bronchoscopy and clinically indicated procedures for suspected infection or respiratory deterioration. For conventional microbiologic testing (CMT), BAL processing was performed by the UPMC clinical microbiology laboratory per institutional protocol, including quantitative bacterial culture, fungal culture, and additional testing (viral PCR, *Pneumocystis jirovecii* PCR, *Aspergillus* galactomannan) as clinically ordered. Excess BAL fluid not required for clinical testing was provided to the research lab where it was aliquoted and stored at −80°C. A total of 33 BAL samples from 19 participants were available for research testing (median 1 per participant, IQR 1–2, range 1–6), and each underwent parallel testing by two metagenomic platforms from the same aliquot.

1. <u>Karius BAL mcfDNA sequencing:</u> BAL mcfDNA sequencing was performed by Karius Inc. (Redwood City, CA) with the Karius Focus test.^***12*,*13***^ As an exploratory research analysis, quantitative cfDNA signals for all detected organisms were provided by Karius Inc. as DNA molecules per microliter (MPM), a proprietary metric of mcfDNA abundance. MPM values were used to characterize longitudinal mcfDNA trajectories.^***19***,***20***^ Samples with no detectable mcfDNA were reported as “No Call.”
2. <u>ONT long-read metagenomics:</u> BAL DNA was extracted following host DNA depletion with a saponin-based protocol, and sequenced on the Oxford Nanopore MinION platform (Mk1c) across three sequencing runs.^***21***,***22***^ Reads were analyzed using the BugSeq bioinformatics pipeline for taxonomic classification.^***23***^ A pre-specified threshold of fewer than 100 total classified microbial reads was applied to define samples with insufficient microbial signal for primary pathogen-level interpretation. Relative abundance was computed as the proportion of classified microbial reads attributable to each taxon within a given sample. Given the overall low yield (see Results), Nanopore findings are reported as a feasibility analysis; an exploratory concordance analysis applied post-hoc thresholds detailed in the Supplementary Methods.

From the same BAL aliquot, host-response biomarkers and microbial markers were measured. Host-response biomarkers included a 10-plex Luminex panel (R&D Systems) comprising angiopoietin-2 (Ang-2), IL-6, IL-8, IL-10, sST2, fractalkine, pentraxin-3, sRAGE, sTNFR1, and procalcitonin;^***24***^ DuoSet ELISA assays (R&D Systems) for syndecan-1 and MMP-7; and a BCA assay for total protein concentration. Lipopolysaccharide (LPS), a microbial cell wall fragment reflecting gram-negative bacterial presence in the airway, was measured separately using a commercially available ELISA (Cusabio, catalog CSB-E09945h).

### Organism classification framework

To enable consistent cross-platform comparisons, all organisms detected by any platform (i.e., conventional culture, Karius BAL mcfDNA sequencing, or Nanopore metagenomics) were assigned to one of three tiers using a pre-defined classification applied uniformly: Tier 1 (pathogens): organisms capable of causing infection in lung transplant recipients and warranting clinical action, including gram-negative rods, gram-positive bacteria, non-*Candida* fungi, mycobacteria, and non-culturable organisms (*Mycoplasma* and *Ureaplasma* species); Tier 2 (commensals/colonization): organisms representing expected airway background microbiota or *Candida* species under universal antifungal prophylaxis; and Tier 3 (uncertain significance): organisms with context-dependent clinical relevance detected at low frequency in this cohort. A small number of Nanopore-detected sequences identified as likely reagent contaminants or organisms with no plausible respiratory pathogen role (bacteriophage, environmental soil organisms, vaginal flora) were removed prior to analysis and are not assigned to any tier. The complete organism classification is provided in Supplementary Table S1.”

### Clinical adjudication and platform concordance classification

Pneumonia and LRTI were adjudicated per adapted National Healthcare Safety Network and International Society for Heart and Lung Transplantation criteria.^25^ Primary Graft Dysfunction (PGD) was adjudicated by two independent reviewers (GDK and MS).

For cross-platform concordance, each BAL episode was assigned to one of four mutually exclusive categories based on genus-level organism matching across metagenomics and culture: (1) concordant pathogen, a Tier 1 pathogen reported by both platforms; (2) metagenomics pathogen / culture missed, metagenomics reported a Tier 1 pathogen while culture was negative, grew normal respiratory flora (NRF) only, or grew yeast only; (3) culture only, culture detected a Tier 1 pathogen while metagenomics returned no Tier 1 pathogen report; and (4) concordant negative, neither platform reported a Tier 1 pathogen, including episodes with concordant yeast or commensal report. This scheme was applied to the BAL mcfDNA vs. culture comparison across all 33 BAL episodes and, as an exploratory analysis, to the Nanopore vs. culture comparison across the 29 episodes with available Nanopore data. For binary concordance, restricted to the first available BAL per patient to avoid pseudoreplication, each episode was classified as positive or negative for each platform and agreement quantified using Cohen’s kappa. Among episodes classified as metagenomics pathogen / culture missed, a predictive report subclassification was applied when the same organism was subsequently confirmed on culture as causing invasive infection in the same participant; lead time was defined as days from first metagenomics detection to culture-confirmed infection.

### Statistical analysis

Descriptive characteristics are provided as numbers (proportions) and medians (inter-quartile range). Spearman rank correlations between BAL biomarkers were computed with Benjamini-Hochberg false discovery rate (FDR) correction. To examine within-subject associations between total mcfDNA burden and BAL biomarkers across serial visits, linear mixed effects models were fitted for biomarkers with available longitudinal measurements across all participants (total protein, MMP-7, Syndecan-1, LPS; N=31–33 episodes, 19 subjects) using log_10_ total MPM (scaled) as a time-varying predictor, visit number as a covariate, and subject as a random intercept (lme4 package, R). All analyses were performed in R version 4.4.4

## RESULTS

### Cohort characteristics and BAL sampling

Between April 2021 and November 2022, 19 lung transplant recipients underwent BAL-based metagenomic testing as part of the KARL prospective cohort. All underwent bilateral transplantation; baseline characteristics are summarized in Table 1. Briefly, median age was 61 years (IQR 53–66), 74% were male, and the most common transplant indications were interstitial lung disease/idiopathic pulmonary fibrosis (37%) and chronic obstructive pulmonary disease/emphysema (32%). Post-transplant morbidity was substantial: 33% required extra-corporeal membrane oxygenation (ECMO) support, 56% underwent tracheostomy, severe PGD (Grade 3) occurred in 26%, and 33% met adjudicated criteria for pneumonia within the first two weeks. One participant died during the observation period. All participants received standard antimicrobial prophylaxis including antifungal agents (Table 1). BAL procedures were performed for surveillance in 24 episodes (73%) and for clinically suspected infection or respiratory deterioration in 9 (27%).

**Table 1.** Baseline characteristics of lung transplant recipients (N = 19)

| Characteristic | N = 19 |
| --- | --- |
| **Demographics** |  |
| Age at transplant, years — median [IQR] | 61 [53, 66] |
| Male sex | 14 (73.7) |
| **Transplant characteristics** |  |
| Bilateral lung transplant | 19 (100) |
| Inpatient at time of transplant<sup>1</sup> | 3 (16.7) |
| **Transplant indication<sup>1</sup>** |  |
| COPD / emphysema | 6 (31.6) |
| ILD / IPF | 7 (36.8) |
| Cystic fibrosis | 1 (5.3) |
| Pulmonary arterial hypertension | 1 (5.3) |
| Other | 4 (21.1) |
| **Induction immunosuppression** |  |
| Alemtuzumab | 7 (36.8) |
| Basiliximab | 12 (63.2) |
| **CMV serostatus** |  |
| **Donor / Recipient<sup>1</sup>** |  |
| Donor– / Recipient– | 5 (27.8) |
| Donor– / Recipient+ | 5 (27.8) |
| Donor+ / Recipient– | 3 (16.7) |
| Donor+ / Recipient+ | 5 (27.8) |
| **Post-transplant clinical outcomes** |  |
| Post-transplant ECMO<sup>1</sup> | 6 (33.3) |
| Tracheostomy<sup>1</sup> | 10 (55.6) |
| Primary graft dysfunction (maximum grade) |  |
| Grade 0 (no edema, PaO<sub>2</sub>/FiO<sub>2</sub> >300) | 1 (5.3) |
| Grade 1 (edema, PaO<sub>2</sub>/FiO<sub>2</sub> >300) | 11 (57.9) |
| Grade 2 (edema, PaO<sub>2</sub>/FiO<sub>2</sub> 200–300) | 2 (10.5) |
| Grade 3 (edema, PaO<sub>2</sub>/FiO<sub>2</sub> <200) | 5 (26.3) |
| Severe PGD (Grade 3) | 5 (26.3) |
| Acute rejection (0–1 month)<sup>1</sup> | 4 (22.2) |
| Pneumonia (first 2 weeks)<sup>1</sup> | 6 (33.3) |
| Death during study period<sup>1</sup> | 1 (5.6) |
| Hospital LOS, days — median [IQR]<sup>1</sup> | 38 [25, 73] |
| **Antimicrobial prophylaxis at 1 month** |  |
| Inhaled amphotericin B<sup>1</sup> | 16 (88.9) |
| Azole antifungal (voriconazole / posaconazole / isavuconazole)<sup>1</sup> | 14 (77.8) |
| Valganciclovir<sup>1</sup> | 13 (72.2) |
| Trimethoprim-sulfamethoxazole<sup>1</sup> | 12 (66.7) |
| Acyclovir<sup>1</sup> | 5 (27.8) |
| **BAL sampling** |  |
| BAL samples per participant — median [IQR] | 1 [1, 2] |

<sup>1</sup> Data missing for KARL\_0031; denominators reflect available data. Percentages calculated on non-missing observations.
<sup>3</sup> ICU LOS calculated as days from transplant date to ICU discharge; 2 participants missing ICU discharge date.
Abbreviations: BAL, bronchoalveolar lavage; CF, cystic fibrosis; CMV, cytomegalovirus; COPD, chronic obstructive pulmonary disease; ECMO, extracorporeal membrane oxygenation; ICU, intensive care unit; ILD, interstitial lung disease; IPF, idiopathic pulmonary fibrosis; IQR, interquartile range; NRF, normal respiratory flora; PAH, pulmonary arterial hypertension; PGD, primary graft dysfunction.

### Technical performance across three platforms

Detection profiles across the three platforms applied to the same BAL specimens are summarized in Figure 1. Microbiologic culture returned a specific bacterial pathogen in 8 of 33 episodes (24%); 7 additional episodes grew yeast only (classified as colonization), and 18 were negative or grew NRF only. The most common pathogens isolated were *Pseudomonas aeruginosa* (n=3), *Burkholderia cepacia* complex (n=2), and methicillin-resistant *Staphylococcus aureus* (n=2).

**Figure 1.**
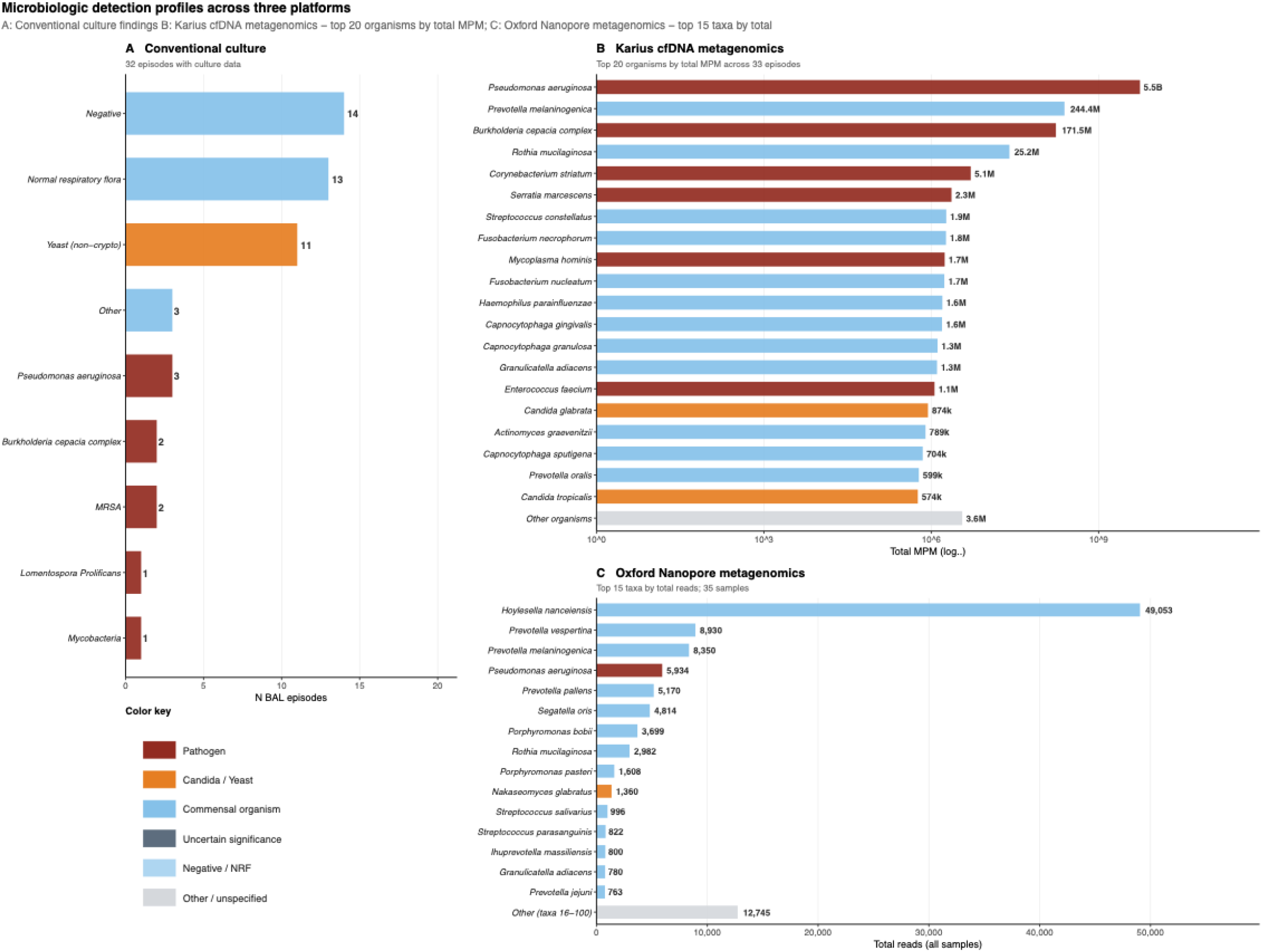
Microbiologic detection profiles across three platforms applied to the same bronchoalveolar lavage specimens. (A) Conventional BAL culture. Number of BAL episodes (N=33) yielding each finding. Light gray = no growth; light blue = normal respiratory flora (NRF); orange = yeast (non-cryptococcal, representing *Candida* spp. classified as colonization under universal antifungal prophylaxis); dark red = specific bacterial pathogen. NRF and yeast were classified as negative or colonization, respectively, for concordance analyses. (B) Karius BAL mcfDNA metagenomics. Top 20 organisms by total cumulative MPM (DNA molecules per microliter, an exploratory research quantitative mcfDNA metric) across all 33 BAL episodes; remaining organisms grouped as Other (light gray). Dark red = Tier 1 adjudicated pathogens, including non-culturable organisms (*Mycoplasma hominis, Ureaplasma parvum*); orange = *Candida* and yeast species (Tier 2, classified as colonization under antifungal prophylaxis); medium blue = oral and airway commensal organisms (Tier 2). X-axis shows log_10_ total MPM. Six episodes returned No Call (no microbial mcfDNA reported above assay threshold). (C) Oxford Nanopore long-read metagenomics. Total classified microbial reads summed across 35 BAL samples for the 15 most abundant taxa; remaining taxa grouped as Other (light gray). Medium blue = oral and upper respiratory commensal taxa; dark red = Tier 1 pathogens (*Pseudomonas aeruginosa* and *Nakaseomyces glabratus*, the teleomorph of *Candida glabrata*). The predominance of commensal taxa and low pathogen-level read depth in most samples precluded primary organism-level concordance analysis; Nanopore findings are reported as a feasibility analysis (Supplementary Figures S1–S2). Colors reflect the unified three-tier organism classification applied throughout. BAL = bronchoalveolar lavage; mcfDNA = microbial cell-free DNA; MPM = molecules per microliter; NRF = normal respiratory flora; Tier 1 = adjudicated pathogen; Tier 2 = commensal/colonization.

Karius BAL mcfDNA reported Tier 1 pathogens in 19 of 33 episodes (58%) and Tier 2 organisms only (*Candida* species or oral commensals) in a further 4 episodes; 6 episodes (18%) returned No Call. The most frequently reported Tier 1 organisms were *P. aeruginosa* (8 episodes), *Enterococcus faecium* (4), *Staphylococcus aureus* (2), and *B. cepacia* complex (2). Non-culturable organisms were reported in 3 episodes: *Mycoplasma hominis* in 2 participants and *Ureaplasma parvum* in 1. Among Tier 2 reports, the most common organisms were *Candida glabrata* (5 episodes), *Candida tropicalis* (5), and oral commensals including *Prevotella melanonigenica* and *Rothia mucilaginosa* (Figure 1B).

Among 35 BAL samples processed by Nanopore metagenomics, 3 (9%) yielded zero classified microbial reads and 26 (74%) fell below the pre-specified threshold of 100 total classified microbial reads (Supplementary Figures S1–S2), leaving 9 samples with sufficient microbial signal for taxonomic analysis. The dominant taxa across these samples were oral commensals (*Hoylesella nanceiensis, Prevotella* species, and *Rothia mucilaginosa*) with *P. aeruginosa* and *Nakaseomyces glabratus* (the teleomorph of *C. glabrata*) the only recognized Tier 1 pathogens in the top 15 taxa by total read abundance (Figure 1C).

### Concordance between metagenomics and conventional BAL culture

Cross-platform concordance across all 33 BAL episodes is summarized in Figure 2A and detailed in Supplementary Table S2. For the primary Karius BAL mcfDNA vs. culture comparison, concordant pathogen report occurred in 6 of 33 episodes (18%), 1 with identical organisms on both platforms and 5 in which BAL mcfDNA confirmed the culture pathogen while identifying additional organisms. In 16 episodes (48%), BAL mcfDNA reported a Tier 1 pathogen while culture was negative, grew NRF only, or grew yeast only. In 2 episodes (6%), culture detected a pathogen while BAL mcfDNA returned no Tier 1 report: *Lomentospora prolificans* (K-9) and yeast with mycobacteria (K-16), both adjudicated as colonization rather than active infection. The remaining 9 episodes (27%) were concordant negative. Binary concordance restricted to the first BAL per patient (N=19) was poor (Cohen’s kappa 0.18, p=0.24).

**Figure 2.**
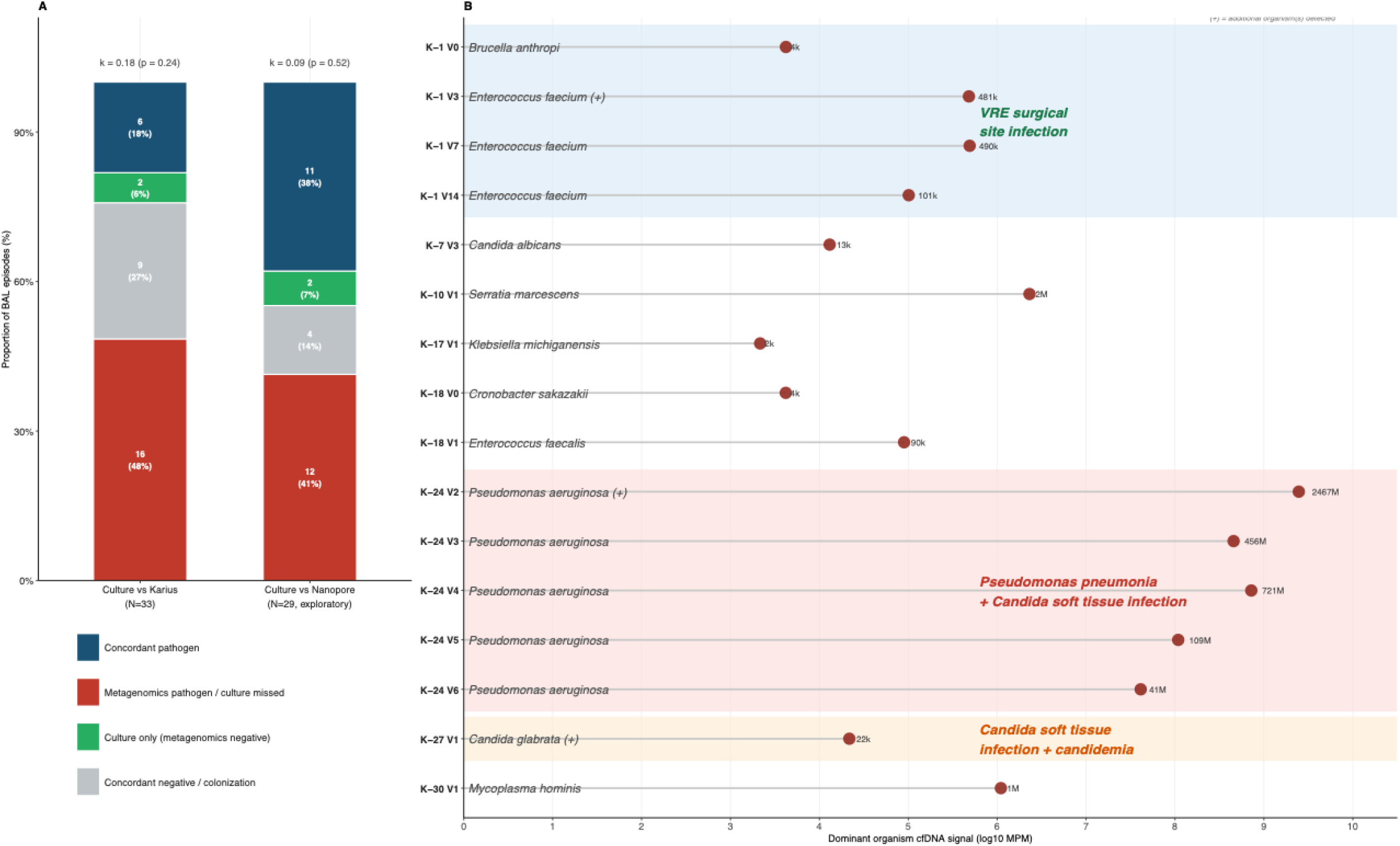
Concordance between metagenomics platforms and conventional BAL culture, and quantitative (by research analysis) BAL mcfDNA signal for Karius-reported pathogens. (A) Distribution of concordance categories across all BAL episodes. Left bar: Karius BAL mcfDNA vs culture (N=33 episodes). Right bar: Nanopore vs culture (N=29 episodes with available Nanopore data, exploratory). Each episode was classified into one of four mutually exclusive categories based on Tier 1 organism report: concordant pathogen = Tier 1 pathogen reported by both platforms at the genus level; metagenomics pathogen / culture missed = metagenomics reported a Tier 1 pathogen while culture was negative, grew NRF only, or grew yeast only; culture only = culture detected a Tier 1 pathogen while metagenomics returned no Tier 1 detection; concordant negative = neither platform reported a Tier 1 pathogen, including episodes with concordant yeast/*Candida* colonization. Cohen’s kappa shown above each bar (first BAL per patient, N=19). (B) Karius BAL mcfDNA reports not recovered by conventional BAL culture (N=16 episodes). Each row represents one BAL episode in which BAL mcfDNA reported one or more Tier 1 pathogens while concurrent culture was negative, grew NRF only, or grew yeast only. X-axis shows the dominant organism mcfDNA signal (log_10_ MPM); (+) indicates additional organisms reported in the same episode. Shaded regions highlight three participants with predictive reports: *Enterococcus faecium* reported across serial BAL visits in Case K-1 before VRE surgical site infection confirmed at POD 28 (blue shading); *Pseudomonas aeruginosa* reported across five consecutive culture-negative BAL visits in Case K-24 before culture-confirmed XDR *Pseudomonas* pneumonia, *Candida glabrata* soft tissue infection, and death at POD 138 (red shading); *C. glabrata* reported in Case K-27 before *C. glabrata* surgical site infection (POD 5) and candidemia (POD 12) (orange shading). BAL = bronchoalveolar lavage; mcfDNA = microbial cell-free DNA; MPM = molecules per million; NRF = normal respiratory flora; No Call = no mcfDNA detected above assay threshold; POD = post-operative day; Tier 1 = adjudicated pathogen per unified organism classification; VRE = vancomycin-resistant *Enterococcus*

As an exploratory analysis, the same four-category scheme was applied to the 29 of 35 Nanopore samples that yielded any classified microbial reads; the 3 samples with zero reads and 3 additional samples without matched culture data were excluded (Figure 2A, right bar). Concordant pathogen detection occurred in 11 of 29 episodes (38%), Nanopore detected a pathogen not recovered by culture in 12 episodes (41%), culture only in 2 (7%), and concordant negative in 4 (14%); binary concordance was similarly poor (Cohen’s kappa 0.09, p=0.52). Given the overall low microbial read yield and post-hoc detection thresholds applied, these findings are interpreted as exploratory.

Among the 16 episodes where Karius BAL mcfDNA reported a Tier 1 pathogen not recovered by culture, the mcfDNA signal (MPM) varied substantially (i.e., from borderline values near background to over 2 billion MPM in the most severely affected participant) with non-culturable organisms (*M. hominis* in 2 participants, K-25 and K-30; *U. parvum* in 1, K-25) (Figure 2B). In eight of these 16 episodes, across 3 participants, the same organism was subsequently confirmed on culture as causing invasive infection 2–46 days later (Figure S3). In Case K-1, *E. faecium* was detected by BAL mcfDNA across three consecutive culture-negative BAL episodes before vanomycin resistant *Enterococcus* (VRE) surgical site infection was confirmed at post-operative day (POD) 28. In Case K-27, *C. glabrata* was reported in a culture-negative BAL specimen on POD 5, the same day as a soft tissue infection, before candidemia was confirmed at POD 12. Case K-24 illustrated a distinct pattern: *P. aeruginosa* and *Candida* species (*C. glabrata, C. tropicalis, Cutaneotrichosporon cutaneum*) were reported by mcfDNA at the first two BAL visits (POD 1–2) while concurrent cultures grew NRF only; from V3 onward, *P. aeruginosa* alone persisted across all remaining BAL visits. At V5 (POD 13), *P. aeruginosa* was also recovered on concurrent culture — the first positive culture in this series — consistent with antibiotic-mediated culture sterilization at earlier visits without true microbiologic eradication. Culture-confirmed XDR *P. aeruginosa* pneumonia recurred repeatedly from POD 63 onward. *C. glabrata* soft tissue infection developed at POD 47, with *C. glabrata* having been reported by mcfDNA at POD 1 and 2 — 45 and 46 days earlier respectively. This participant died at POD 138. Longitudinal mcfDNA trajectories for these three cases, including quantitative MPM across serial BAL visits and timing of culture-confirmed clinical events, are shown in Supplementary Figure S3.

### Perioperative pathogen origins

To explore whether early Karius BAL mcfDNA reports reflected donor-derived transmission, recipient-derived persistence, or novel post-transplant acquisition, we compared the first available post-transplant Karius BAL result (V1 in 15 participants; V2 in 2) against donor sterility cultures and recipient explant cultures (Supplementary Table S3). Among 17 participants with early BAL mcfDNA data, organism-level comparisons identified three distinct patterns. Potential donor-derived transmission was identified in 1 participant (K-10), in whom *Serratia marcescens* was reported by BAL mcfDNA at V1 while concurrent BAL culture was negative and the recipient explant was sterile, but the donor sterility culture had grown *Serratia*. Recipient-derived persistence was identified in 1 participant (K-26), in whom *B. cepacia* complex was concordantly reported by both BAL mcfDNA and explant culture on V1, consistent with persistence of the recipient’s own organism into the transplanted allograft. In 6 participants, BAL mcfDNA reported organisms absent from both donor and recipient cultures, suggesting novel early post-transplant acquisition; these included *P. aeruginosa* in K-24 and *C. glabrata* in K-27, both of which subsequently caused invasive infections, and *M. hominis* in K-30.

### BAL host-response biomarkers

As an exploratory analysis, we examined correlations between BAL biomarkers and quantitative Karius BAL mcfDNA signal (total MPM) across all 33 episodes (Figure 3). Total MPM showed significant positive correlations with multiple inflammatory biomarkers after FDR correction: total protein (ρ=0.64, p_adj<0.001, N=33), IL-6 (ρ=0.72, p_adj=0.027, N=15), Fractalkine (ρ=0.66, p_adj=0.038, N=15), IL-8 (ρ=0.62, p_adj=0.043, N=15), and sST2 (ρ=0.60, p_adj=0.043, N=15).

**Figure 3.**
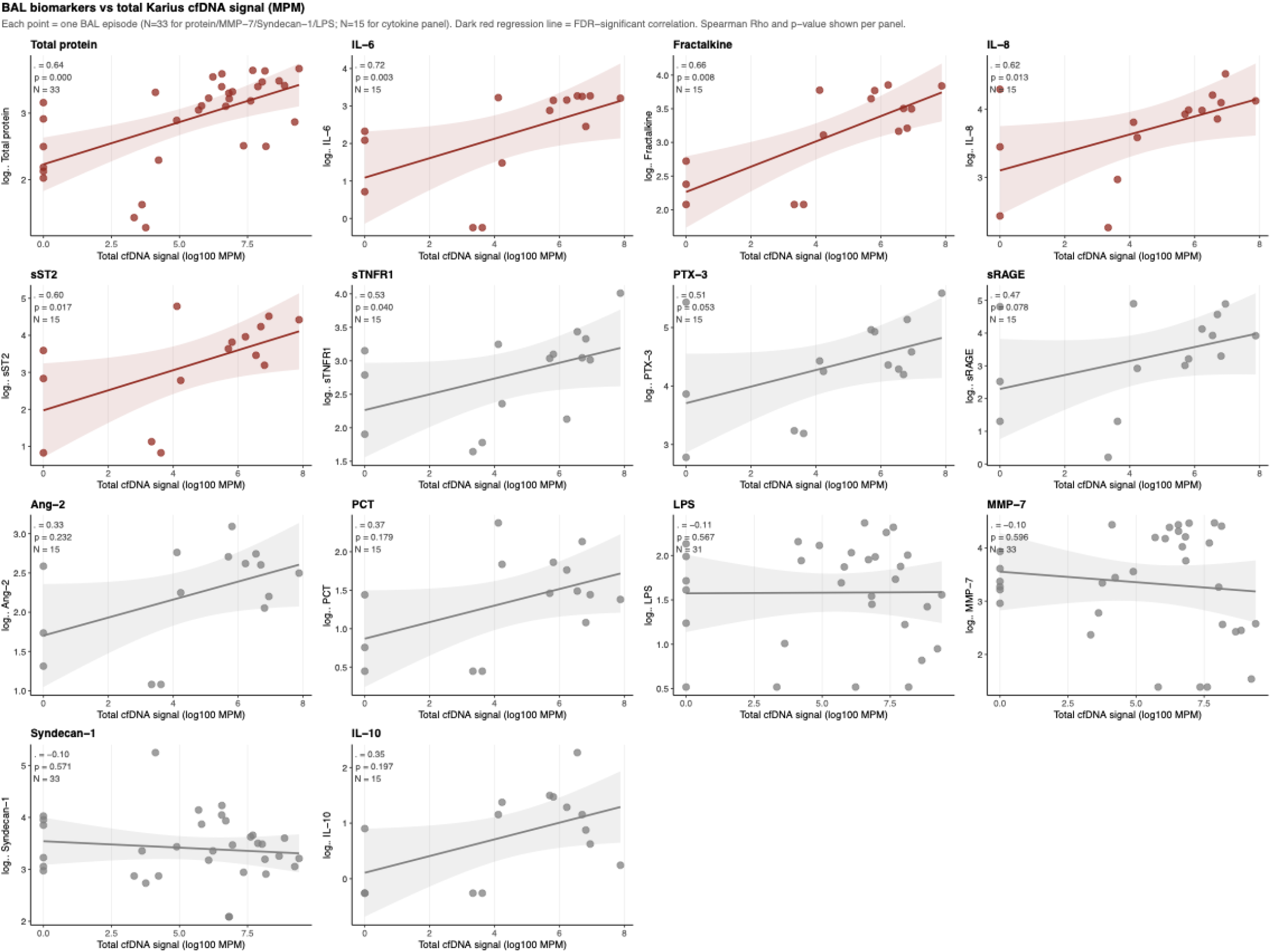
BAL biomarkers vs total Karius BAL mcfDNA signal. Each point represents one BAL episode. X-axis shows log_10_ total MPM, a quantitative metric of cumulative microbial mcfDNA burden per sample, determined by research analysis. Y-axis shows log_10_ biomarker concentration (raw, unadjusted). Total protein, MMP-7, Syndecan-1, and LPS were available in all 33 episodes; cytokine panel (IL-6, Fractalkine, IL-8, sST2, sTNFR1, PTX-3, sRAGE, Ang-2, PCT, IL-10) was available in 15 episodes. Dark red regression lines indicate FDR-significant correlations (Benjamini-Hochberg); gray lines indicate nominal associations. Spearman ρ and unadjusted p-value shown per panel. BAL = bronchoalveolar lavage; mcfDNA = microbial cell-free DNA; MPM = DNA molecules per microliter

In illustrative longitudinal analyses of two predictive report cases (K-1 and K-24), BAL inflammatory biomarkers, including sST2, IL-6, Fractalkine, and PTX-3, showed parallel co-elevation with *E. faecium* cfDNA signal across serial visits in Case K-1 prior to VRE surgical site infection, as well as persistent elevation of Syndecan-1 and MMP-7 with *P. aeruginosa* mcfDNA report, providing case-level support for the cross-sectional MPM-inflammation correlations observed across the cohort (Figure S4).

In linear mixed effects models examining within-subject associations across serial BAL visits, total BAL protein showed a significant positive association with total MPM after FDR correction (β=0.39 per SD increase in log_10_ MPM, p_adj=0.018, N=33 episodes, 19 subjects, Figure S5), indicating that visits with higher mcfDNA burden had higher alveolar protein content within the same patient. MMP-7, Syndecan-1, and LPS showed no within-subject association with total MPM. In an exploratory analysis, BAL biomarkers showed no significant correlation with Nanopore total classified reads or Tier 1 pathogen reads in the 29 samples with available Nanopore data (all FDR p>0.05; data not shown), consistent with the overall low microbial read yield limiting the Nanopore signal.

## DISCUSSION

In this prospective cohort of lung transplant recipients undergoing serial bronchoscopy in the immediate post-transplant period, BAL metagenomics using two distinct sequencing approaches revealed a microbiologic landscape substantially broader than that captured by conventional culture alone. Karius BAL mcfDNA metagenomic sequencing reported Tier 1 pathogens in 19 of 33 episodes (58%) compared with 8 of 33 episodes (24%) by conventional culture, identified non-culturable organisms invisible to standard microbiologic methods, and reported pathogens in culture-negative specimens that subsequently caused invasive infections in 3 of 19 participants. Nanopore sequencing, applied as a feasibility analysis of multiplexed specimens, was limited by the high proportion of contaminating human DNA in BAL specimens, a well-recognized challenge of whole cell metagenomic sequencing in this specimen type,^26^ resulting in insufficient microbial read depth in most samples. Organism-level concordance between Karius and conventional culture was limited, with true pathogen agreement in only 6 of 32 evaluable episodes, reflecting the fundamentally distinct detection mechanisms of the two platforms: conventional culture requires viable organisms capable of *ex vivo* growth, while mcfDNA metagenomics detects nucleic acid fragments shed into the airway lining fluid regardless of organism viability, culturability, or concurrent antimicrobial exposure.

The most clinically consequential finding was the report of pathogens in culture-negative BAL specimens that preceded invasive infections in 3 of 19 recipients by 2 to 46 days. These predictive reports spanned organisms (i.e., *E. faecium, C. glabrata*, and *P. aeruginosa*) with fundamentally different clinical implications and occurred under conditions of universal prophylaxis that rendered conventional culture insensitive, highlighting a window of diagnostic opportunity that CMT cannot access. Prior studies of plasma mcfDNA metagenomic sequencing in immunocompromised patients have demonstrated similar predictive capacity,^27^ but BAL-based detection localizes the signal to the LRT and may be particularly informative in lung transplant recipients, in whom the airway is the primary site of early pathogen acquisition and invasion.

The K-24 case illustrates a mechanistically distinct pattern that deserves separate consideration. Rather than only representing a predictive report of a new pathogen, persistent *Pseudomonas* mcfDNA across six consecutive culture-negative BAL episodes (with MPM values exceeding 10□ molecules per microliter at peak) likely reflected ongoing endobronchial pathogen burden despite apparent culture sterilization under antibiotic pressure. The *Candida* species reported at the first two visits, 45–46 days before culture-confirmed *C. glabrata* soft tissue infection, adds a second layer to this case: an early warning signal for a pathogen that was present but not recovered by concurrent culture, likely suppressed by antifungal prophylaxis. This phenomenon has been described in chronic airway infection^28^ but has not previously been documented by serial mcfDNA surveillance in the post-transplant setting. The subsequent development of multiresistant *Pseudomonas* pneumonia in this participant raises the hypothesis that persistent mcfDNA detection despite negative cultures may signal impending treatment failure rather than microbiologic clearance. Whether mcfDNA trajectories (i.e., rising, stable, or falling across serial BAL episodes) could inform decisions about antimicrobial de-escalation or treatment intensification warrants prospective investigation.

The report of non-culturable organisms (e.g., *Mycoplasma hominis* and *Ureaplasma parvum*) in culture-negative BAL specimens adds a category of detection inaccessible to CMT. Both organisms have been increasingly recognized as causes of respiratory failure and tracheobronchitis in lung transplant recipients,^29^ and their identification requires either dedicated PCR or culture-independent methods. The perioperative source analysis further illustrates the unique potential of mcfDNA metagenomic sequencing to attribute early pathogen detections to donor transmission, recipient-derived persistence, or novel acquisition, a distinction that CMT cannot reliably make and that has direct implications for antimicrobial stewardship and infection prevention.

Oxford Nanopore sequencing, applied with saponin-based host depletion and multiplexed across runs of approximately 12 samples for cost efficiency,^17,21,30^ yielded insufficient microbial read depth in the majority of BAL specimens, precluding organism-level concordance analysis. BAL presents particular challenges for metagenomic sequencing relative to sputum or endotracheal aspirate samples: the BAL procedure introduces substantial dilution of airway lining fluid, and the high proportion of alveolar macrophages and epithelial cells generates a heavy human DNA background even after depletion.^26^ We and others have successfully generated high-yield microbial metagenomics from sputum and endotracheal aspirate samples using the MinION platform,^20,22,31^ suggesting that the low yield observed here reflects specimen-type constraints rather than an intrinsic limitation of nanopore sequencing for respiratory pathogen detection. Alternative approaches, including deeper sequencing per sample without multiplexing, adaptive sampling to enrich microbial reads in real time, or optimized host depletion protocols tailored to BAL, may improve yield sufficiently for clinical application.

BAL host-response biomarkers were profiled to characterize the alveolar inflammatory milieu in relation to metagenomic findings. Total BAL mcfDNA burden showed significant positive cross-sectional correlations with multiple alveolar inflammatory mediators after FDR correction, including total protein, IL-6, Fractalkine, IL-8, and sST2. Within-subject linear mixed effects models further demonstrated a significant positive association between total MPM and BAL total protein across serial visits, consistent with a relationship between mcfDNA burden and alveolar barrier disruption or inflammatory exudate. These findings suggest that quantitative mcfDNA signal may carry biological information beyond conventional diagnostic labeling and offer an opportunity to further characterize host-microbiota interactions in the immediate post-transplant period in future studies. Nanopore total reads did not correlate significantly with any biomarker, consistent with the low overall microbial read yield. The cytokine panel was available in a subset of episodes from the first enrollment cohort only, limiting cytokine-MPM interpretation to cross-sectional observations.

Several limitations of this study deserve acknowledgment. The cohort is single-center and small, limiting statistical power for biomarker analyses and precluding definitive conclusions about the frequency of predictive reports across the broader lung transplant population. The study was observational, and Karius results were not available to the clinical team in real time; we therefore cannot assess whether BAL mcfDNA-guided preemptive treatment would have altered outcomes in the three predictive cases. Our analyses focused on infectious outcomes and early post-transplant complications including PGD; the relationship between early LRT microbiota and longer-term allograft outcomes, including acute cellular rejection and chronic lung allograft dysfunction was not examined and represents an important direction for future investigation.^32,33^ Finally, the cost and infrastructure requirements of mcfDNA metagenomics currently limit its deployment to specialized centers, and the optimal timing, frequency, and clinical decision thresholds for mcfDNA-guided surveillance bronchoscopy remain to be defined.

In conclusion, BAL mcfDNA metagenomics identified clinically relevant pathogens not recovered by conventional culture in the immediate post-transplant period, including organisms preceding invasive infection by days to weeks and non-culturable organisms that are hard to detect with CMT. These findings support prospective evaluation of mcfDNA-based surveillance in lung transplant recipients, with particular attention to the clinical significance of persistent pathogen detection in the context of apparent culture sterilization and the potential for integrating quantitative mcfDNA trajectories with host biomarkers to refine the interpretation of metagenomic findings and guide treatment decisions.

## Data Availability

All data produced in the present study are available upon reasonable request to the authors

## Acknowledgements

The authors thank Sarah Park, Matt Smollin, and Tim Blauwkamp (Karius Inc.) for conducting microbial cell-free DNA sequencing and for their input in the interpretation of results. The authors thank Kelly Friday, MD, Christopher Musgrove, MD, Angela Pisarra, and Ellen Morell (University of Pittsburgh Medical Center) for their contributions to participant enrollment and clinical data abstraction. We are grateful to the patients and families who participated in this observational research during a challenging period in their care. We thank the nursing and respiratory therapy staff at UPMC Presbyterian for their dedication to participant recruitment and research sample acquisition.

## List of Abbreviations

Ang-2: angiopoietin-2
BAL: bronchoalveolar lavage
BCA: bicinchoninic acid assay
CF: cystic fibrosis
CLAD: chronic lung allograft dysfunction
CMT: conventional microbiologic testing
CMV: cytomegalovirus
COPD: chronic obstructive pulmonary disease
ECMO: extracorporeal membrane oxygenation
ELISA: enzyme-linked immunosorbent assay
FDR: false discovery rate
ICU: intensive care unit
ILD: interstitial lung disease
IL-6: interleukin-6
IL-8: interleukin-8
IL-10: interleukin-10
IPF: idiopathic pulmonary fibrosis
IQR: interquartile range
IRB: Institutional Review Board
KARL: Karius After Recent Lung transplant
LOS: length of stay
LPS: lipopolysaccharide
LRTI: lower respiratory tract infection
LRT: lower respiratory tract
lmcfDNA: microbial cell-free DNA
MMP-7: matrix metalloproteinase-7
MPM: DNA molecules per microliter
MRSA: methicillin-resistant *Staphylococcus aureus*
NRF: normal respiratory flora
ONT: Oxford Nanopore Technology
PAH: pulmonary arterial hypertension
PCR: polymerase chain reaction
PCT: procalcitonin
PGD: primary graft dysfunction
POD: post-operative day
PTX-3: pentraxin-3
sRAGE: soluble receptor for advanced glycation end-products
sST2: soluble suppression of tumorigenicity-2
sTNFR1: soluble tumor necrosis factor receptor 1
UPMC: University of Pittsburgh Medical Center
VRE: vancomycin-resistant *Enterococcus*
XDR: extensively drug-resistant

