## supplement for "Bronchoalveolar Lavage Metagenomic Sequencing in the Early Post-Lung Transplantation Period: A Pilot Comparison with Microbiologic Culture"

**Extended Methods: Platform concordance classification**

**Overview**

Concordance between Karius bronchoalveolar lavage (BAL) microbial cell-free DNA (mcfDNA) metagenomics, Oxford Nanopore metagenomics, and conventional culture was assessed using a unified four-category scheme applied to all three pairwise comparisons. All organisms reported by any platform were first assigned to one of three tiers using a pre-defined classification applied uniformly (see Supplementary Table S1): Tier 1 (adjudicated pathogen), Tier 2 (commensal/colonization), and Tier 3 (uncertain significance). Only Tier 1 organisms were used for concordance classification; Tier 2 and Tier 3 organisms were not counted as pathogens for this purpose.

**Culture result classification**

Prior to concordance assessment, each culture result was assigned to one of three states using a strict priority hierarchy:

*Pathogen*: at least one Tier 1 organism was identified. Co-occurrence of NRF or yeast alongside a Tier 1 organism did not alter the pathogen classification.

*Colonization*: yeast was the only discrete finding, with or without concurrent NRF, and no Tier 1 organism was identified. This classification reflects the clinical context of universal antifungal prophylaxis, under which *Candida* detected in BAL represents airway colonization rather than invasive infection.

*Negative*: no growth, NRF only, or any combination thereof without yeast or a Tier 1 organism. NRF was classified as negative rather than colonization because it does not represent a discretely identified organism and carries no pathogen-specific diagnostic information.

The hierarchy was applied in strict priority order: pathogen > colonization > negative.

**Metagenomics result classification**

Karius BAL mcfDNA and Nanopore results were each assigned to one of two states for concordance:

*Pathogen reported*: at least one Tier 1 organism was reported. For Karius, this required a pathogen report with non-*Candida* Tier 1 organism; *Candida* reports were treated as colonization consistent with the antifungal prophylaxis context. For Nanopore, pathogen detection required meeting all three pre-specified thresholds (≥50 total classified reads, ≥10 Tier 1 pathogen reads, ≥5% relative abundance; see Supplementary Methods: Nanopore thresholds).

*No pathogen reported*: No Call, commensal-only report, or *Candida*-only report by Karius; or failure to meet pathogen reporting thresholds by Nanopore.

Four-category concordance scheme

Each BAL episode was assigned to one of four mutually exclusive categories:

*(1) Concordant pathogen*: a Tier 1 pathogen reported by both metagenomics and culture at the genus level.

*(2) Metagenomics pathogen / culture missed*: metagenomics reported a Tier 1 pathogen while culture was negative, grew NRF only, or grew yeast only. This category includes non-culturable organisms (*Mycoplasma*, *Ureaplasma*) detectable only by culture-independent methods. Among episodes in this category, a predictive report subclassification was applied when the same organism was subsequently confirmed on conventional culture as causing invasive infection in the same participant (see below).

*(3) Culture only*: culture detected a Tier 1 pathogen while metagenomics returned no Tier 1 detection. For Karius, this corresponds to a No Call result in the presence of a culture pathogen.

*(4) Concordant negative*: neither platform reported a Tier 1 pathogen, including episodes with concordant yeast/*Candida* colonization on both platforms and episodes where Karius BAL mcfDNA reported commensal organisms only.

For binary concordance restricted to the first BAL per patient, each episode was classified as positive (Tier 1 pathogen detected) or negative for each platform independently and agreement quantified using Cohen's kappa.

**Genus-level organism matching**

For concordance at the genus level, culture results and metagenomics reports were mapped to standardized genus identifiers. Methicillin sensitive and resistant *Staphylococcus* *aureus* were mapped to *Staphylococcus*; yeast (non-cryptococcal, culture) was mapped to *Candida*; *Cronobacter sakazakii* was mapped to *Cronobacter*; *Burkholderia cepacia* complex was mapped to *Burkholderia*. Genus-level rather than species-level matching was used to account for differential taxonomic resolution between platforms.

**Predictive report subclassification**

Among episodes classified as metagenomics pathogen / culture missed, a subclassification of predictive report was applied post hoc when all three criteria were met: (a) metagenomics detected a Tier 1 non-*Candida* pathogen while concurrent culture was negative or NRF; (b) the same organism at the genus level was subsequently confirmed on conventional culture from any clinical specimen from the same participant; and (c) the culture confirmation was associated with a clinically diagnosed invasive infection adjudicated from the electronic medical record. Lead time was defined as days from the first qualifying metagenomics report to the first culture-confirmed invasive infection with the same organism.

**Nanopore post-hoc detection thresholds**

Given the overall low microbial read yield, a pre-specified threshold of fewer than 100 total classified reads was used to define samples with insufficient microbial signal for primary interpretation. For the exploratory concordance analysis, pathogen detection by Nanopore required all three of the following: ≥50 total classified microbial reads per sample, ≥10 reads classified as a Tier 1 pathogen, and ≥5% relative abundance of Tier 1 pathogen reads. These thresholds were applied post hoc to minimize false-positive pathogen calls from index hopping between multiplexed samples and reagent contamination at very low read counts.

**Quantitative mcfDNA signal (MPM)**

Quantitative mcfDNA signal was provided by Karius Inc. as DNA molecules per microliter (MPM), a proprietary metric of mcfDNA abundance generated by research analysis. MPM values were available for all 33 Karius samples for reported organisms; samples with No Call results were assigned MPM = 1 (log₁₀ = 0) to indicate absence of reportable signal. Organism-specific MPM was computed by summing MPM across all reported organisms within a tier (Tier 1 pathogen MPM, *Candida*-specific MPM). Spearman correlations between log₁₀ MPM and log₁₀ BAL biomarker concentrations were computed across all available episodes. Within-subject associations were tested using linear mixed effects models (lme4 package, R) with log₁₀ total MPM as a time-varying predictor, visit number as a covariate, and subject as a random intercept, restricted to core biomarkers available across all 33 episodes.

**Supplementary Table S1. Unified organism classification framework applied uniformly across all three detection platforms**

| **Tier** | **Organisms reported in this cohort** | | **Platforms** |
| --- | --- | --- | --- |
| **Tier 1 Pathogens** | *Acinetobacter baumannii complex; Aspergillus spp.; Burkholderia cepacia complex; Burkholderia multivorans; Corynebacterium durum; Corynebacterium striatum; Cronobacter sakazakii; Cryptococcus spp.; Enterobacter spp.; Enterococcus faecalis; Enterococcus faecium; Escherichia coli; Klebsiella michiganensis; Klebsiella spp.; Lomentospora prolificans; Mucorales; Mycobacteria spp.; Mycoplasma hominis; Nocardia spp.; Pseudomonas aeruginosa; Serratia marcescens; Staphylococcus aureus; Staphylococcus aureus (MRSA/MSSA); Stenotrophomonas maltophilia; Ureaplasma parvum* | Culture; Karius; Nanopore | |
| **Tier 2 Commensal / colonization** | *Actinomyces graevenitzii; Actinomyces marseillensis; Actinomyces naeslundii; Actinomyces oris; Aggregatibacter segnis; Alloprevotella tannerae; Arachnia propionica; Bacteroides fragilis; Bifidobacterium longum; Campylobacter concisus; Campylobacter gracilis; Campylobacter showae; Candida albicans; Candida dubliniensis; Candida glabrata (syn. Nakaseomyces glabratus); Candida parapsilosis; Candida tropicalis; Capnocytophaga gingivalis; Capnocytophaga granulosa; Capnocytophaga leadbetteri; Capnocytophaga ochracea; Capnocytophaga sputigena; Cardiobacterium hominis; Cutaneotrichosporon cutaneum; Cutibacterium acnes; Dolosigranulum pigrum; Eikenella corrodens; Fusobacterium necrophorum; Fusobacterium nucleatum; Gemella haemolysans; Gemella sanguinis; Granulicatella adiacens; Haemophilus haemolyticus; Haemophilus parahaemolyticus; Haemophilus parainfluenzae; Haemophilus paraphrohaemolyticus; Hoylesella enoeca; Hoylesella nanceiensis; Ihuprevotella massiliensis; Lacticaseibacillus rhamnosus; Lancefieldella parvula; Lancefieldella rimae; Megasphaera micronuciformis; Mogibacterium diversum; Neisseria elongata; Neisseria perflava; Neisseria sicca; Neisseria subflava; Normal respiratory flora (NRF); Olsenella profusa; Olsenella uli; Peptostreptococcus anaerobius; Peptostreptococcus stomatis; Phocaeicola vulgatus; Porphyromonas bobii; Porphyromonas catoniae; Porphyromonas endodontalis; Porphyromonas gingivalis; Porphyromonas pasteri; Prevotella bivia; Prevotella buccae; Prevotella conceptionensis; Prevotella denticola; Prevotella fusca; Prevotella histicola; Prevotella ihumii; Prevotella intermedia; Prevotella jejuni; Prevotella melaninogenica; Prevotella nigrescens; Prevotella oralis; Prevotella pallens; Prevotella scopos; Prevotella veroralis; Prevotella vespertina; Rothia aeria; Rothia dentocariosa; Rothia mucilaginosa; Scardovia wiggsiae; Schaalia dentiphila; Schaalia odontolytica; Segatella baroniae; Segatella maculosa; Segatella oris; Segatella oulorum; Segatella salivae; Slackia exigua; Solobacterium moorei; Stomatobaculum longum; Streptococcus anginosus; Streptococcus australis; Streptococcus constellatus; Streptococcus gordonii; Streptococcus infantis; Streptococcus mitis; Streptococcus oralis; Streptococcus parasanguinis; Streptococcus peroris; Streptococcus pseudopneumoniae; Streptococcus salivarius; Streptococcus sanguinis; Streptococcus timonensis; Tannerella serpentiformis; Treponema vincentii; Veillonella atypica; Veillonella dispar; Veillonella nakazawae; Veillonella parvula; Veillonella rogosae; Yeast (non-cryptococcal, culture)* | Culture; Karius; Nanopore | |
| **Tier 3 Uncertain significance** | *Actinomyces israelii; Brucella anthropi (syn. Ochrobactrum anthropi); Corynebacterium propinquum* | Karius; Nanopore | |
| **Excluded Non-organism / contaminant** | *Fannyhessea vaginae; Isoptericola variabilis; Lacnuvirus LcNu* | Nanopore | |

Abbreviations: NRF, normal respiratory flora; MRSA, methicillin-resistant *Staphylococcus aureus*; MSSA, methicillin-susceptible *Staphylococcus aureus*. Tier 1 includes non-culturable organisms (*Mycoplasma hominis*, *Ureaplasma parvum*) reported only by culture-independent methods. Tier 2 includes all *Candida* species classified as colonization under universal antifungal prophylaxis, as well as oral and upper respiratory commensal flora. Tier 3 organisms (*Actinomyces israelii*, *Brucella anthropi*, *Corynebacterium propinquum*) have context-dependent clinical significance and were reported at low frequency; they are not counted as pathogens in concordance analyses. Excluded organisms (bacteriophage, environmental soil organism, vaginal flora) detected at very low read counts by Nanopore metagenomics were removed from all analyses as likely contaminants.

**Supplementary Figure S1.** Nanopore metagenomics read depth and microbial richness by study visit. Panel A: Total classified microbial reads per sample on log^10^ scale by visit group (V0, V1, V2, V3+). Dashed line indicates 100-read pre-specified interpretability threshold. Red filled triangles indicate three samples with zero microbial reads. Boxes show IQR; horizontal line = median; whiskers = 1.5×IQR. Panel B: Number of taxa detected per sample by visit group.

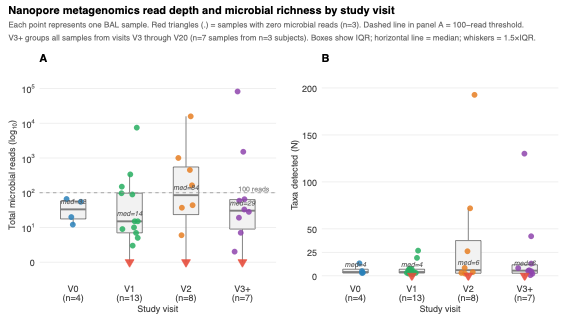

**Supplementary Figure S2.** Nanopore relative abundance by subject. Stacked barplot showing within-sample relative abundance for the top 15 taxa and Other across 33 BAL samples, faceted by subject (N=19). Each bar represents one BAL sample and sums to 100%. Samples with fewer than 100 microbial reads are marked with an asterisk (*); samples with zero microbial reads are marked with 0.

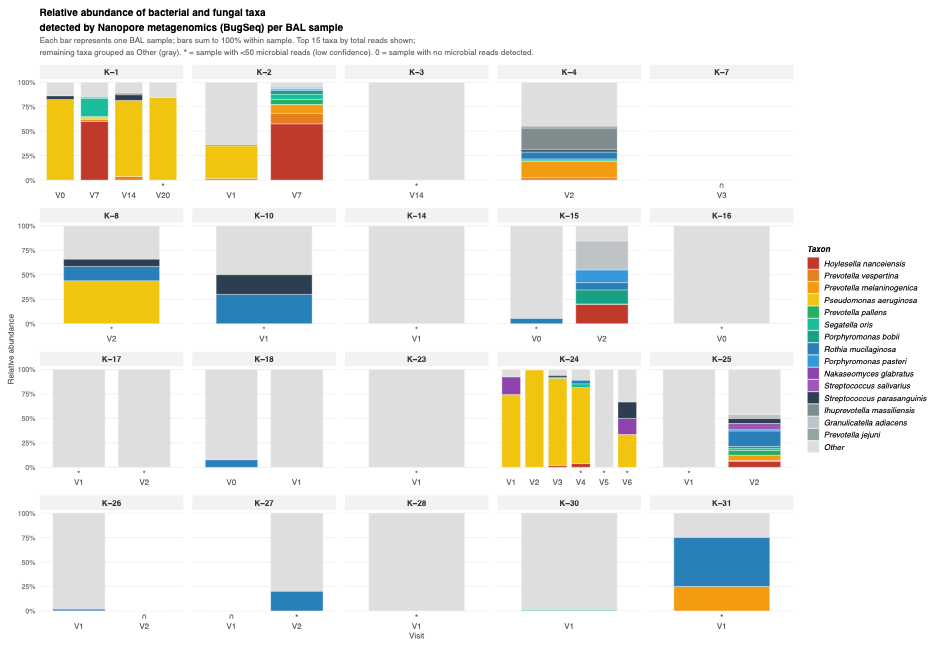

**Supplementary Table S2. Organism-level concordance between Karius BAL mcfDNA metagenomics and conventional BAL culture — organized by subject.**

Each row represents one BAL episode; shading groups episodes by subject. All 19 subjects and 33 BAL episodes are included. Tier 1 (adjudicated pathogen) organisms reported by Karius BAL mcfDNA are listed in the Karius column; concordant organisms (same genus detected by both platforms) are shown in bold blue italic. Tier 2 (commensal/colonization) organisms, including Candida species and oral commensals, are shown in small gray italic below the Tier 1 results. Organism-level agreement categories: Organism-concordant pathogen = same Tier 1 genus reported by both platforms; Karius expanded = concordant Tier 1 organism plus Karius identified additional organisms not recovered by culture; Karius pathogen / culture colonization = culture grew yeast only (colonization under antifungal prophylaxis), Karius reported a distinct Tier 1 pathogen; Karius-only predictive = culture negative or NRF, Karius reported Tier 1 pathogen, same organism subsequently confirmed on culture as causing invasive infection; Karius-only (no infection) = culture negative, Karius reported Tier 1 pathogen, no subsequent infection identified; Culture only (Karius No Call) = culture grew a Tier 1 pathogen, Karius reported no mcfDNA above threshold; Concordant colonization = yeast on culture, Candida species reported by Karius (both classified as Tier 2 colonization); Concordant negative = neither platform reported a Tier 1 pathogen, including episodes with No Call. BAL = bronchoalveolar lavage; mcfDNA = microbial cell-free DNA; NRF = normal respiratory flora (classified as negative); No Call = no mcfDNA detected above the assay threshold (valid negative result, not a technical failure); POD = post-operative day; SSI = surgical site infection; VRE = vancomycin-resistant Enterococcus; XDR = extensively drug-resistant

| **Case** | **Visit** | **Indication** | **Conventional culture result** | **Karius mcfDNA (**Tier 1 (pathogen-level) organisms listed; Tier 2 (commensal/colonization) organisms shown in gray italic below) | **Organism-level agreement** | **Clinical interpretation (per case)** |
| --- | --- | --- | --- | --- | --- | --- |
| **K-1** | V0 | Surveillance | *Yeast (non-crypto); NRF* | *Brucella anthropi (Ochrobactrum anthropi)* *Haemophilus parainfluenzae; Rothia mucilaginosa; Streptococcus pseudopneumoniae* | **Karius pathogen / culture colonization** | Four BAL episodes. V0: culture grew yeast + NRF; Karius reported Brucella/Ochrobactrum (GNR), a platform-discordant finding with no clinical correlate. V3 and V14: culture negative; Karius reported Enterococcus faecium — the VRE SSI pathogen confirmed on wound drainage culture ~3 weeks after V3. V7: culture grew yeast (colonization); Karius reported E. faecium while culture failed to recover it. E. faecium reported by Karius across 3 visits (V3, V7, V14) in the absence of positive cultures, constituting a predictive report. Outcome: VRE (E. faecium) surgical site infection; alive. |
|  | V3 | Clin. indicated | *Negative* | *Candida dubliniensis* *Enterococcus faecium* *Actinomyces graevenitzii; Granulicatella adiacens; Haemophilus parainfluenzae; Rothia mucilaginosa; Slackia exigua* | **Karius-only predictive** |  |
|  | V7 | Surveillance | *Yeast (non-crypto); NRF* | *Enterococcus faecium* *Bifidobacterium longum; Capnocytophaga gingivalis; Rothia mucilaginosa* | **Karius pathogen / culture colonization** |  |
|  | V14 | Clin. indicated | *Negative* | *Bifidobacterium longum; Capnocytophaga gingivalis; Eikenella corrodens; Rothia mucilaginosa* | **Karius-only predictive** |  |
| **K-2** | V2 | Surveillance | *Yeast (non-crypto); NRF* | *Actinomyces graevenitzii; Capnocytophaga granulosa; Fusobacterium necrophorum; Fusobacterium nucleatum; Haemophilus parainfluenzae; Prevotella melaninogenica; Rothia mucilaginosa; Streptococcus anginosus; Streptococcus constellatus* | **Concordant colonization** | Two BAL episodes. V2: culture grew yeast + NRF; Karius reported commensals only — concordant colonization. V7: culture negative; Karius Cat3 only — concordant negative. No clinical infection identified. |
|  | V7 | Surveillance | *Negative* | *Actinomyces graevenitzii; Capnocytophaga granulosa; Fusobacterium necrophorum; Fusobacterium nucleatum; Prevotella melaninogenica; Rothia mucilaginosa; Streptococcus constellatus* | **Concordant negative** |  |
| **K-7** | V3 | Surveillance | *Negative* | *Candida albicans* | **Karius-only (no infection)** | One BAL episode. V3: culture negative; Karius reported Candida albicans (classified as colonization). No subsequent Candida infection identified. Patient alive; on treatment for Nocardia isolated from explanted lung (unrelated to transplanted allograft). |
| **K-8** | V2 | Surveillance | *Yeast (non-crypto)*  *Pseudomonas aeruginosa* | *Actinomyces israelii* *Candida albicans* ***Pseudomonas aeruginosa*** *Actinomyces oris; Capnocytophaga ochracea; Fusobacterium nucleatum; Rothia mucilaginosa* | **Karius expanded (concordant + additional)** | One BAL episode. Karius reported Pseudomonas (concordant with clinical chart), plus Actinomyces israelii and Candida albicans additionally. Classified as Karius expanded. Outcome: LRTI caused by Pseudomonas, treated with cefepime; resolved. |
| **K-9** | V14 | Clin. indicated | *Lomentospora prolificans (known colonizer); bacterial culture negative* | *No Call* *(no mcfDNA reported)* | **Culture only (Karius No Call)** | One BAL episode. V14: culture grew Lomentospora prolificans, a known pre-transplant colonizer adjudicated as not causing active infection. Karius returned No Call (no mcfDNA reported). No clinical infection identified. |
| **K-10** | V1 | Clin. indicated | *Negative; NRF* | *Serratia marcescens* *Rothia mucilaginosa* | **Karius-only (no infection)** | One BAL episode. V1: culture negative + NRF (treated as negative); Karius reported Serratia marcescens. Note: donor sterility culture also grew Serratia — possible donor-derived detection. No subsequent Serratia infection confirmed. Patient developed pneumatosis coli later (unrelated). |
| **K-14** | V1 | Surveillance | *Negative* | *No Call* *(no mcfDNA reported)* | **Concordant negative (No Call)** | One BAL episode. V1: culture negative; Karius No Call. Both platforms negative. Bronchial anastomotic complications without positive microbiology. No clinical infection. |
| **K-15** | V1 | Surveillance | *Negative* | *Streptococcus pseudopneumoniae* | **Concordant negative** | Two BAL episodes. V1 and V2: culture negative; Karius reported Cat3 commensals only at both visits. Both platforms negative for pathogens. No clinical infection. |
|  | V2 | Surveillance | *Negative* | *Capnocytophaga gingivalis; Fusobacterium nucleatum; Granulicatella adiacens; Haemophilus parainfluenzae; Rothia mucilaginosa; Schaalia odontolytica* | **Concordant negative** |  |
| **K-16** | V0 | Surveillance | *Yeast (non-crypto); Mycobacteria; NRF* | *No Call* *(no mcfDNA reported)* | **Culture only (Karius No Call)** | One BAL episode. V0: culture grew yeast + mycobacteria + NRF; Karius No Call. Respiratory symptoms attributed to acute rejection (not infection); empiric antibiotics stopped after biopsy. No clinical infection. |
| **K-17** | V1 | Surveillance | *Negative* | *Klebsiella michiganensis* | **Karius-only (no infection)** | One BAL episode. V1: culture negative; Karius reported Klebsiella michiganensis. No subsequent Klebsiella infection documented. Likely early post-transplant acquisition. Patient alive. |
| **K-18** | V0 | Surveillance | *Yeast (non-crypto)* | *Cronobacter sakazakii (Enterobacter sakazakii)* | **Karius pathogen / culture colonization** | Two BAL episodes. V0: culture grew yeast only; Karius reported Cronobacter sakazakii (GNR) not recovered on culture — platform-discordant. V1: culture grew yeast + NRF; Karius reported Enterococcus faecalis and broad flora. Patient had proven Ureaplasma tracheobronchitis (PCR-confirmed); improved with levofloxacin (Enterococcus coverage). |
|  | V1 | Clin. indicated | *Yeast (non-crypto); NRF* | *Enterococcus faecalis* *Actinomyces oris; Bacteroides fragilis; Campylobacter gracilis; Fusobacterium nucleatum; Granulicatella adiacens; Rothia mucilaginosa; Streptococcus constellatus* | **Karius pathogen / culture colonization** |  |
| **K-23** | V1 | Surveillance | *Negative* | *No Call* *(no mcfDNA reported)* | **Concordant negative (No Call)** | One BAL episode. V1: culture negative; Karius No Call. Both platforms negative. No clinical infection. |
| **K-24** | V1 | Clin. indicated | *NRF (negative)* | *Candida glabrata* *Candida tropicalis* *Cutaneotrichosporon cutaneum* *Pseudomonas aeruginosa* *Bifidobacterium longum* | **Karius-only predictive** | Six BAL episodes. V1–V4: culture grew NRF only at all visits; Karius reported Pseudomonas aeruginosa from V1 onward — constituting predictive reports. Culture-confirmed Pseudomonas was first recovered on a clinical BAL at POD 9 (V5). V6: culture NRF. XDR Pseudomonas pneumonia subsequently confirmed per ISHLT criteria with recurrent culture positivity from POD 63 onward. Candida glabrata soft tissue infection at POD 47; death at POD 138.. |
|  | V2 | Surveillance | *NRF (negative)* | *Candida glabrata* *Candida tropicalis* *Cutaneotrichosporon cutaneum* *Pseudomonas aeruginosa* | **Karius-only predictive** |  |
|  | V3 | Surveillance | *NRF (negative)* | *Pseudomonas aeruginosa* | **Karius-only predictive** |  |
|  | V4 | Surveillance | *NRF (negative)* | *Pseudomonas aeruginosa* | **Karius-only predictive** |  |
|  | V5 | Surveillance | *Pseudomonas aeruginosa; NRF* | ***Pseudomonas aeruginosa*** | **Karius expanded (concordant + additional)** |  |
|  | V6 | Surveillance | *NRF (negative)* | *Pseudomonas aeruginosa* | **Karius-only predictive** |  |
| **K-25** | V1 | Surveillance | *MRSA* | *Mycoplasma hominis* ***Staphylococcus aureus*** *Actinomyces oris; Capnocytophaga gingivalis; Capnocytophaga granulosa; Capnocytophaga ochracea; Capnocytophaga sputigena; Corynebacterium propinquum; Corynebacterium striatum; Dolosigranulum pigrum; Fusobacterium nucleatum; Granulicatella adiacens; Haemophilus haemolyticus; Haemophilus parainfluenzae; Peptostreptococcus anaerobius; Prevotella melaninogenica; Rothia mucilaginosa; Schaalia odontolytica; Slackia exigua* | **Karius expanded (concordant + additional)** | Two BAL episodes. V1: culture MRSA; Karius reported S. aureus (concordant with MRSA) plus non-culturable Mycoplasma hominis — Karius expanded. V2: Karius reported Pseudomonas, S. aureus, and Ureaplasma parvum. Outcome: proven Pseudomonas + MRSA pneumonia (ISHLT criteria); chronic respiratory failure, tracheostomy-dependent. |
|  | V2 | Other | Pseudomonas aeruginosa \| MRSA | *Pseudomonas aeruginosa* *Staphylococcus aureus* *Ureaplasma parvum* | **Karius expanded (concordant + additional)** |  |
| **K-26** | V1 | Surveillance | *Yeast (non-crypto); Burkholderia cepacia complex* | ***Burkholderia cepacia complex*** | **Organism-concordant pathogen** | Two BAL episodes. V1: culture grew yeast + Burkholderia cepacia complex; Karius reported Burkholderia — organism-concordant. V2: same culture pattern; Karius additionally reported Candida albicans, C. glabrata, and Enterococcus faecium — Karius expanded. Known pre-transplant Burkholderia colonization (CF patient) persisting into allograft. No active infection identified. |
|  | V2 | Surveillance | *Yeast (non-crypto); Burkholderia cepacia complex* | ***Burkholderia cepacia complex*** *Candida albicans* *Candida glabrata* *Enterococcus faecium* | **Karius expanded (concordant + additional)** |  |
| **K-27** | V1 | Clin. indicated | *Negative* | *Candida glabrata* *Candida tropicalis*  *Capnocytophaga granulosa; Granulicatella adiacens; Prevotella buccae* | **Karius-only predictive** | Two BAL episodes. V1: culture negative; Karius reported Candida glabrata and C. tropicalis — Karius-only predictive. C. glabrata surgical site infection confirmed POD 5; C. glabrata fungemia POD 12. V2: culture negative; Karius No Call — concordant negative. Alive; chronic airway colonization. |
|  | V2 | Clin. indicated | *Negative* | *No Call* *(no mcfDNA reported)* | **Concordant negative (No Call)** |  |
| **K-28** | V1 | Surveillance | *Negative* | *No Call* *(no mcfDNA reported)* | **Concordant negative (No Call)** | One BAL episode. V1: culture negative; Karius No Call. Both platforms negative. Patient later developed Enterobacter pneumonia (culture-confirmed, treated with ciprofloxacin) — not anticipated by this surveillance BAL. |
| **K-30** | V1 | Surveillance | *Yeast (non-crypto); NRF* | *Mycoplasma hominis* *Corynebacterium striatum;*  *Prevotella oralis; Streptococcus constellatus* | **Karius pathogen / culture colonization** | One BAL episode. V1: culture grew yeast + NRF; Karius reported Mycoplasma hominis — a non-culturable organism not recoverable on standard media. Platform-discordant (culture colonization vs. Karius bacterial report). No Mycoplasma infection clinically confirmed. |
| **K-31** | V1 | Other | *Yeast (non-crypto)* | *Candida dubliniensis* *Candida glabrata* *Candida tropicalis* *Prevotella melaninogenica; Rothia mucilaginosa* | **Concordant colonization** | One BAL episode. V1: culture detected yeast; Karius reported Candida complex. Concordance for colonization. |

**Supplementary Table S3. Comparison of donor sterility culture and recipient explant culture with earliest available post-transplant Karius cfDNA BAL result (V1 preferred; V2 if V1 unavailable)**

All 19 subjects in the Karius cohort are included. Karius results reflect the first available post-transplant BAL sample (V1 in 15 subjects; V2 in 2 subjects; no V1/V2 available in 4 subjects). Donor sterility cultures are from BAL obtained at organ procurement. Recipient explant cultures are from the native lung BAL obtained at the time of transplant surgery (from Supplementary Table S2). Putative source classification: Donor-derived = same genus detected in donor sterility culture and Karius Cat2+, with recipient explant negative or non-concordant; Recipient-derived = same genus in recipient explant and Karius Cat2+; Novel acquisition = organism absent from both donor sterility and recipient explant cultures. Candida concordance is noted but treated as colonization throughout given universal antifungal prophylaxis. No Call = no mcfDNA reported above assay threshold (valid negative result). NRF = normal respiratory flora; GNR = gram-negative rods; POD = post-operative day; SSI = surgical site infection.

| **Case** | **Donor sterility culture** | **Recipient explant culture** | **Visit** | **Karius cfDNA (earliest post-transplant)** | **Putative source** | **Clinical interpretation** |
| --- | --- | --- | --- | --- | --- | --- |
| **Potential donor-derived detection — Karius concordant with donor sterility culture (n=3)** | | | | | | |
| **K-10** | *Pseudomonas aeruginosa; Serratia spp.* | *Negative* | V1 | *Serratia marcescens* | **Donor-concordant (Serratia)** | Karius reported Serratia concordant with donor sterility culture; concurrent BAL culture negative. Recipient explant negative — organism absent from recipient prior to transplant. Possible donor-derived transmission. |
| **K-25** | *MSSA* | *MRSA* | V1 | *Staphylococcus aureus; Mycoplasma hominis* | **Donor- or recipient- concordant (Staph)** | S. aureus concordant with both donor (MSSA) and recipient explant (MRSA) — source ambiguous. Karius additionally reported non-culturable Mycoplasma hominis. Proven Pseudomonas + MRSA pneumonia. |
| **K-8** | *Candida* | *Pseudomonas aeruginosa* | V2 | *Candida albicans; Pseudomonas aeruginosa; Actinomyces israelii* | **Donor-concordant (Candida, colonization)** | Candida concordant with donor sterility culture (colonization level). Pseudomonas concordant with recipient explant. Karius additionally reported Actinomyces. LRTI caused by Pseudomonas (treated, resolved). |
| **Recipient-derived persistence — Karius concordant with recipient explant culture (n=1)** | | | | | | |
| **K-26** | *MRSA; Citrobacter; Candida* | *Burkholderia cepacia complex* | V1 | *Burkholderia cepacia complex* | **Recipient-concordant (Burkholderia)** | Burkholderia concordant with recipient explant — known pre-transplant CF colonization persisting into allograft. Donor MRSA and Candida not reported by Karius at V1. |
| **Novel post-transplant acquisition — organism absent from both donor and recipient cultures (n=6)** | | | | | | |
| **K-24** | *MSSA* | *NRF* | V1 | *Pseudomonas aeruginosa; Candida glabrata; C. tropicalis; Cutaneotrichosporon* | **Novel acquisition** | Pseudomonas and Candida absent from both donor and recipient cultures. Early post-transplant acquisition. Pseudomonas caused fatal XDR pneumonia; Candida glabrata caused soft tissue infection (POD 47). |
| **K-27** | *MSSA; Enterobacter spp.* | *Negative* | V1 | *Candida glabrata; Candida tropicalis* | **Novel acquisition** | Candida absent from donor and recipient cultures. Early post-transplant acquisition. C. glabrata surgical site infection (POD 5) and fungemia (POD 12). |
| **K-17** | *MSSA* | *Negative* | V1 | *Klebsiella michiganensis* | **Novel acquisition** | Klebsiella absent from donor and recipient cultures. Early post-transplant acquisition. No subsequent Klebsiella infection confirmed. |
| **K-18** | *Candida* | *Negative* | V1 | *Enterococcus faecalis* | **Novel acquisition** | Enterococcus absent from donor and recipient cultures. Patient improved with levofloxacin (Enterococcus coverage). |
| **K-30** | *MSSA; Candida* | *MSSA* | V1 | *Mycoplasma hominis* | **Novel acquisition (non-culturable)** | Mycoplasma hominis not reported in donor or recipient cultures — non-culturable organism invisible to standard microbiologic methods. No Mycoplasma infection clinically confirmed. |
| **K-31** | *Negative* | *Not available* | V1 | *Candida dubliniensis; Candida glabrata; Candida tropicalis* | **Novel acquisition (indeterminate)** | Donor culture negative. Recipient data unavailable. Clinical significance indeterminate. |
| **Karius negative or No Call at earliest available visit (n=5)** | | | | | | |
| **K-2** | *MRSA* | *Negative* | V2 | *Commensal only* | **Karius negative** | No pathogen reported by Karius at V2. |
| **K-14** | *Acinetobacter baumannii* | *Negative* | V1 | *No Call* | **No Call** | Donor had Acinetobacter; Karius returned No Call — no mcfDNA reported. |
| **K-15** | *Candida* | *NRF* | V1 | *Commensal only* | **Karius negative** | Donor Candida not reported by Karius. |
| **K-23** | *MSSA* | *Negative* | V1 | *No Call* | **No Call** | No mcfDNA reported. |
| **K-28** | *MSSA* | *Negative* | V1 | *No Call* | **No Call** | No mcfDNA reported. Enterobacter pneumonia developed later. |
| **No V1 or V2 Karius sample available (n=4)** | | | | | | |
| **K-1** | *MSSA* | *NRF* | — | *No V1/V2 sample (V0 only; earliest post-transplant Karius at V3)* | **No sample** | VRE SSI reported by Karius at V3/V7/V14. |
| **K-7** | *Enterobacter; Veillonella; Campylobacter* | *Nocardia (explanted lung)* | — | *No V1/V2 sample (earliest at V3)* | **No sample** | V3 Karius reported Candida only; no donor-concordant detection. |
| **K-9** | *MRSA; MSSA* | *NRF* | — | *No V1/V2 sample (earliest at V14)* | **No sample** | Lomentospora colonization at V14; Karius No Call. |
| **K-16** | *Serratia; Enterobacter* | *Negative* | — | *V0 only (No Call — insufficient reads)* | **No sample (V0 No Call)** | Donor had GNR; Karius V0 No Call. Symptoms attributed to rejection. |

**Figure S3. Longitudinal BAL mcfDNA signal trajectories in three predictive report cases.**

Each point represents one Karius BAL sample; y-axis shows log₁₀ MPM for each reported organism. Lines connect organisms reported at multiple visits. Filled circles = Tier 1 adjudicated pathogen; open circles = Tier 2 organism (Candida species or oral commensal). Vertical dashed lines indicate culture-confirmed clinical events. X-axis shows days post-transplant (POD) independently for each case.

(A) Case K-1. Enterococcus faecium was reported by Karius across three consecutive BAL visits (POD 5–13) in the absence of concurrent positive cultures, preceding VRE surgical site infection confirmed at POD 28 (green dashed line). Oral commensal organisms (open circles) were reported at all visits.

(B) Case K-24. Pseudomonas aeruginosa was reported at high mcfDNA signal across six consecutive BAL visits (POD 1–14) while concurrent cultures were negative after the first visit; culture-confirmed Pseudomonas was first reported at POD 9 (dark red dashed line). Candida glabrata and C. tropicalis were reported at low signal at early visits, preceding C. glabrata soft tissue infection at POD 47 (orange dashed line). Multiple subsequent culture-confirmed Pseudomonas episodes occurred through POD 133; death at POD 138.

(C) Case K-27. Candida glabrata and C. tropicalis were detected at POD 5, the same day as a C. glabrata soft tissue infection (orange dashed line) and 7 days before C. glabrata candidemia (red dashed line).

BAL = bronchoalveolar lavage; mcfDNA = microbial cell-free DNA; MPM = DNA molecules per microliter; POD = post-operative day; Tier 1 = adjudicated pathogen; Tier 2 = commensal/colonization; VRE = vancomycin-resistant Enterococcus.

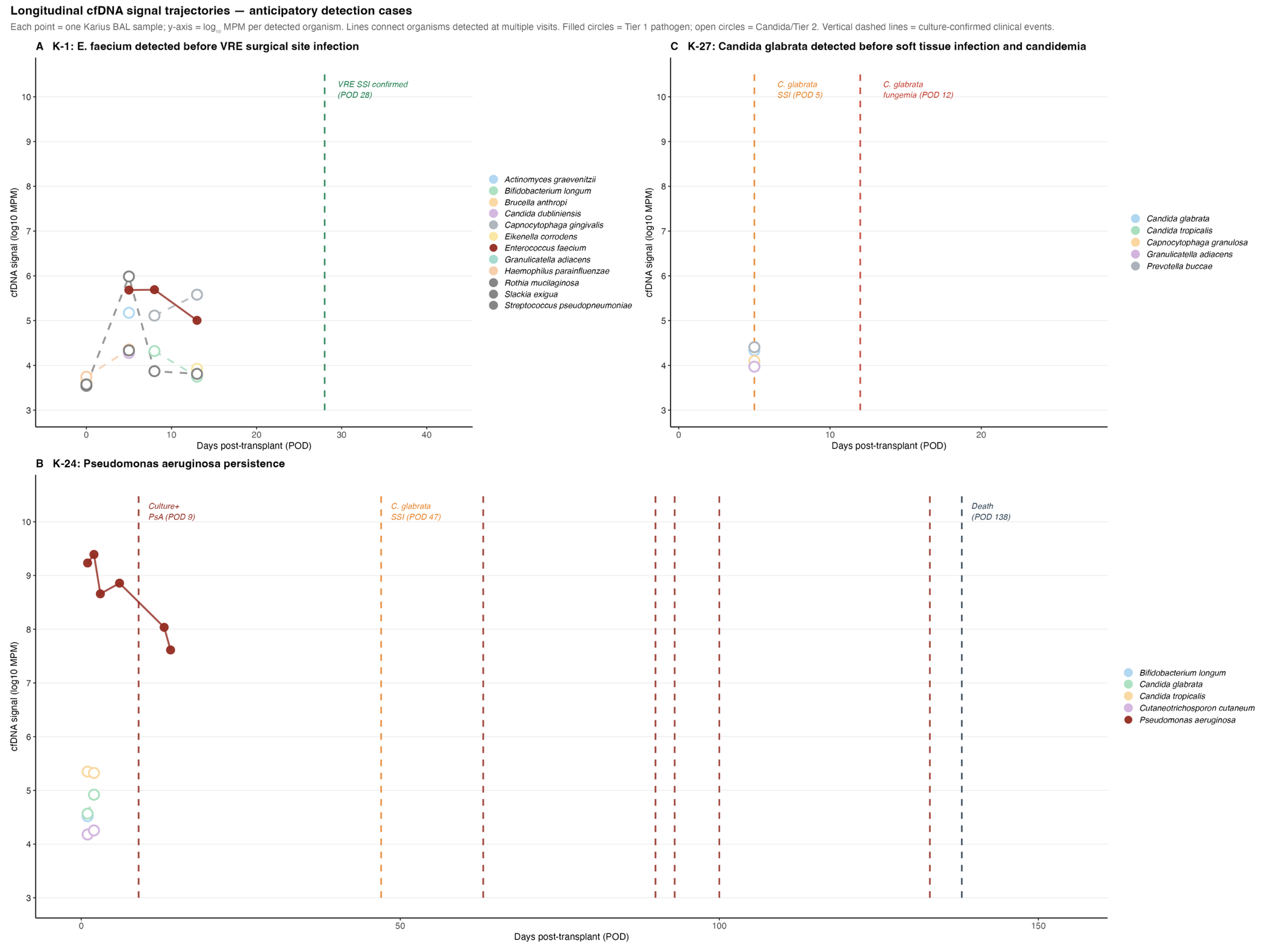

**Figure S4. Longitudinal BAL mcfDNA signal and BAL biomarker trajectories in two predictive report cases.** Each panel shows log₁₀-transformed values across serial BAL visits plotted by days post-transplant (POD). (A) Case K-1: *Enterococcus faecium* cfDNA signal (MPM) and concurrent BAL biomarkers across four visits (POD 0–13). *E. faecium* was absent at V0 (MPM set to 1; log₁₀=0) and detectable from V3 onward, with VRE surgical site infection confirmed at POD 28 (green dashed line). (B) Case K-24: *Pseudomonas aeruginosa* cfDNA signal and core BAL biomarkers across six visits (POD 1–14); cytokine panel was not available for this participant. Culture-confirmed *Pseudomonas* at POD 9 and *Candida glabrata* soft tissue infection at POD 47 are indicated by dashed lines. Y-axes show raw unadjusted biomarker concentrations. BAL = bronchoalveolar lavage; mcfDNA = microbial cell-free DNA; MPM = DNA molecules per microliter; VRE= vancomycin resistant *Enterococcus*.

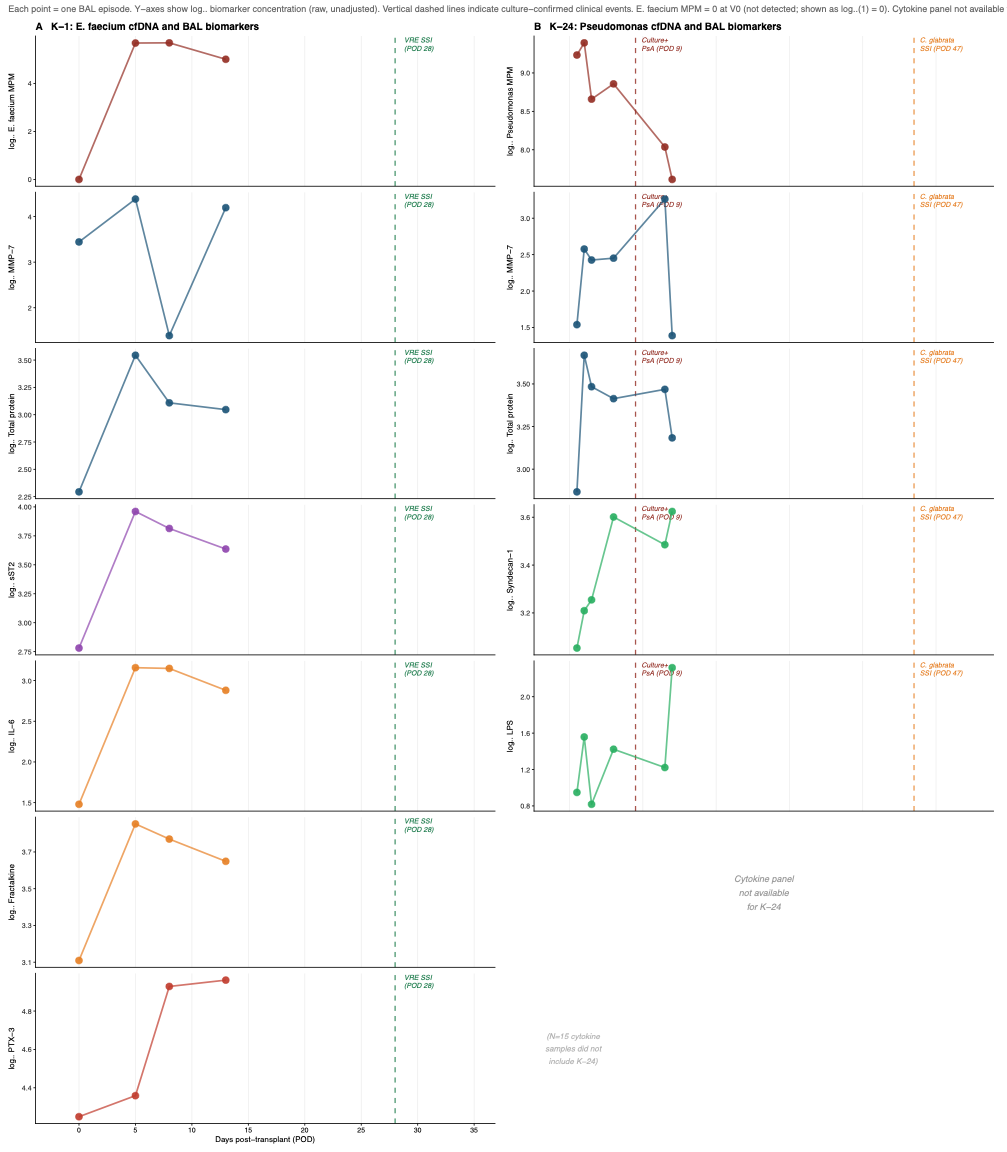

**Figure S5. Within-subject association between total Karius BAL mcfDNA signal and core BAL biomarkers.** Linear mixed effects model: log₁₀(biomarker) ~ log₁₀ total MPM (scaled) + visit number + (1|Subject). Points show β coefficient ± 95% CI per 1 SD increase in log₁₀ total MPM. Dark red = FDR<0.05; gray = non-significant. N=31–33 episodes across 19 subjects. Cytokine panel excluded due to enrollment cohort batch effect. BAL = bronchoalveolar lavage; MPM = molecules per microliter; FDR = Benjamini-Hochberg.

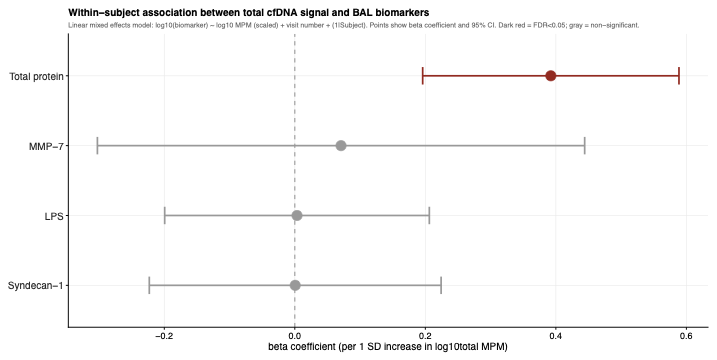
